# Automation improves repeatability of retinal oximetry measurements

**DOI:** 10.1101/2021.08.25.21262154

**Authors:** Robert Arnar Karlsson, Olof Birna Olafsdottir, Soumaya Belhadj, Vedis Helgadottir, Thorunn Scheving Eliasdottir, Einar Stefansson, Sveinn Hakon Hardarson

## Abstract

**Purpose:** Retinal oximetry is a technique based on spectrophotometry where images are analyzed with software capable of calculating vessel oxygen saturation and vessel diameter. In this study, the effect of automation of measurements of retinal vessel oxygen saturation and vessel diameter is explored.

**Methods:** Until now, operators have had to choose each vessel segment to be measured explicitly. A new, automatic version of the software automatically selects the vessels once the operator defines a measurement area.

Five operators analyzed image pairs from the right eye of 23 healthy subjects with semiautomated retinal oximetry analysis software, Oxymap Analyzer (v2.5.1), and an automated version (v3.0). Inter- and intra-operator variability was investigated using the intraclass correlation coefficient (ICC) between oxygen saturation measurements of vessel segments in the same area of the retina.

**Results:** For manual saturation measurements, the inter-rater ICC was 0.80 for arterioles and venules. For automated saturation measurements, the inter-rater ICC was 0.97 for arterioles and 0.96 for venules. For manual diameter measurements, the inter-rater ICC was 0.71 for arterioles and venules. For automated diameter measurements the inter-rater ICC was 0.97 for arterioles and 0.95 for venules. The inter-rater ICCs were different (*p <* 0.01) between the semiautomated and automated version in all instances.

**Conclusion:** Automated measurements of retinal oximetry values are more repeatable compared to measurements where vessels are selected manually.

## Introduction

Fundus photographs are widely used to document various ocular diseases such as diabetic retinopathy [1], glaucoma [2], retinopathy of prematurity [3] and retinal vein occlusion [4]. The retinal blood vessels are visible through the optics of the eye and can be imaged with a fundus camera. Changes in retinal vessel oxygen saturation have been found in diabetic retinopathy, glaucoma, age-related macular degeneration, and other common diseases affecting the retina [5–8] as well as systemic and brain disease [9].

Measurements of retinal vessel oxygen saturation were first attempted in 1959 [10], and several groups have experimented with different approaches since then (for review, see [11–14]).

Currently, there are three commercial retinal oximeters available. One of them is the Oxymap T1 which simultaneously acquires two images of the same area of the fundus at two different wavelengths of light. The two spectral images are processed by specialized software, Oxymap Analyzer. However, the Oxymap T1 is limited by the fact that measurements are semiautomated. As a result, users are required to choose each vessel segment being measured and classify it as an arteriole or venule. Manual input can lead to operator-induced variability in the measurement results. In addition, the analysis process is time-consuming and tedious. Therefore, new, automatic software has been developed which automatically detects the vessels, classifies them as arterioles or venules, and selects the vessels once the operator defines a measurement area. This paper evaluates the effect of automation on the repeatability of measurements of retinal vessel oxygen saturation and diameter.

## Methods

Five consecutive images of the right eye were taken with the optic nerve head located in different part of the image. The second and fifth image had the optic nerve head located in the center and those two images were used for analysis. All images were acquired in a dark room with the aiming light set to lowest setting. The flash intensity was set to 50 Ws. The time between flashes (images) never exceeded 30 seconds. The right eye from 23 healthy volunteers (aged between twenty and thirty years) was used for measuring.

Two optic-nerve-centered images were acquired with an interval of no more than two minutes, from the right eye of 23 healthy volunteers aged between twenty and thirty years.

Collection and analysis of images were done with the approval of The Icelandic Data Protection Authority with the informed consent of patients or volunteers and following the tenets of the Declaration of Helsinki.

### Retinal oximetry

The retinal oximeter (Oxymap T1; Oxymap ehf., Reykjavik, Iceland) consists of an image splitter and two digital cameras (Insight IN1800; Diagnostic Instruments Inc, Michigan, USA) attached to a fundus camera (Topcon TRC-50DX; Topcon Corporation, Tokyo, Japan). The device simultaneously acquires two images of the same area of the fundus at two different wavelengths of light, one sensitive to oxyhemoglobin (600 nm) and one isosbestic (570 nm), where oxyhemoglobin and hemoglobin absorb the same amount of light. The images are 1200 × 1600 pixels and cover a 50°field of the retina.

The light absorbance of a solution can be described in terms of optical density (OD) at wavelength *λ*. Optical density is defined as *OD*_*λ*_ = log(*I*_0*λ*_*/I*_*λ*_) where *I*_0*λ*_ is the intensity of the incoming light and *I*_*λ*_ is the intensity of the light after it has interacted with and been absorbed to some degree by the solution in question. Here *I*_0*λ*_ and *I*_*λ*_, are estimated from brightness values chosen respectively from reflected light inside vessels, and the perivascular background in the fundus images [7].

If one of the wavelengths is sensitive to oxyhemoglobin (e.g. 600nm) and one is isosbestic (e.g. 570nm), the ratio of the optical densities ODR = *OD*_570*nm*_*/OD*_600*nm*_ at these two wavelengths will have an inverse and approximately linear relationship to the oxygen saturation (SatO2) [15]. Studies have demonstrated an artifact in the measurements caused by different diameters of the vessels [15–17] making it necessary to add a correction term.

The oxygen saturation is then calculated using *Sat*02 = *a · ODR* + *b* + *c · D* + *k* where *D* is the diameter of the vessel in pixels, and the parameters *a, b, c* and *k* can be calibrated based on measurements of arteries and veins of healthy volunteers and assuming values from saturation measurements performed in a study with a calibrated device [17, 18].

For version 2.5 of the software the calibration parameters were set to *a* = *−*1.28, *b* = 1.24, *c* = 0.0097 and *k* = *−*0.14. For version 3.0 of the software the calibration parameters were set to *a* = *−*1.28, *b* = 1.24, *c* = 0.0095 and *k* = *−*0.107.

In version 2.5 *I*_*λ*_ is selected as the darkest pixel value along the cross-section of a vessel for each wavelength, meaning that saturation values are only calculated for points close to the center of a vessel (unless there is a central reflex from the vessel). For version 3.0, all points within a vessel are used to calculate saturation values.

### Analysis of oximetry images

Images were analyzed with a previously validated semiautomatic software [19–21] (Oxymap Analyzer v.2.5.1, referred to as version 2.5) and new, automatic software (referred to as version 3.0). Figure 1 shows screenshots from both versions where the measurement area is defined.

**Fig 1.**
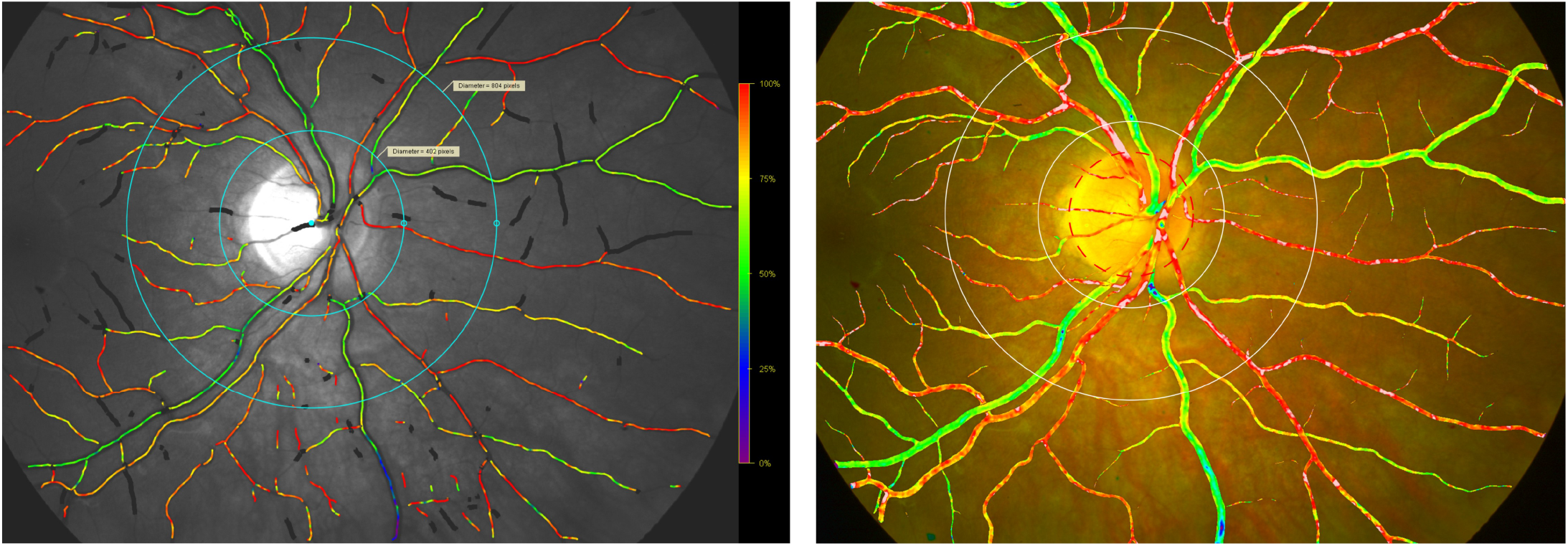
Screenshots of the same image analyzed with version 2.5 (left) version 3.0 (right). Pseudocolor maps are generated automatically. Colors indicate the retinal vessel oxygen saturation. In healthy individuals arteries are normally colored orange to red (approximately 90% - 100% saturation). Veins are colored green to yellow-green (approximately 50% - 60% saturation). Both software versions detect the retinal vessels automatically and select measurement points inside and outside of the retina for automatic calculation of the retinal vessel oxygen saturation.

The first step of the analysis process for both versions is to select the optic nerve head in the image manually. The measurement area is then defined as the area between an inner circle with a diameter 1.5 times the optic nerve diameter and an outer circle with a diameter 3.0 times the diameter of the optic nerve diameter (Figure 1). The measurement area is defined to avoid reflection of light from the retinal nerve fiber layer around the optic nerve to make the measurements more accurate. For version 2.5, vessel segments with a diameter of 8 pixels or greater between the two circles were manually selected according to a standardized protocol (Oxymap ehf. protocol from November 2013) and classified as arteries or venules by the operator. The saturation and diameter values for each vessel segment would then be exported to an external application for further analysis. Mean values are calculated from all measured arterioles and all measured venules. In version 3.0, the software would automatically summarize the saturation and diameter values for arterioles and venules once the operator had selected the optic nerve head in the image. Median values of all measurement points are used, for arterioles and venules separately.

### Statistical analysis

The software versions are compared in several ways. The standard deviation of measured values is compared using an F-test in arterioles and venules for both saturation and diameter. Intraclass correlation coefficients (ICCs), and their 95% confidence intervals, are used to assess the intra (*ICC*_*intra*_.) and inter-operator (*ICC*_*inter*_) reliability using a two-way random factorial absolute agreement ANOVA model [22]. When interpreting reliability using the ICC scores, “values less than 0.5 are indicative of poor reliability, values between 0.5 and 0.75 indicate moderate reliability, values between 0.75 and 0.9 indicate good reliability, and values greater than 0.90 indicate excellent reliability” [22]. While the ICC method is widely used when comparing methods or operators, it has been pointed out that it can, under some circumstances, misleadingly indicate good performance [23]. We, therefore, analyze the repeatability of measurements with the two versions using the 95% limits of agreement method (LoA) [24]. The repeated measures are for all operators, separately for oxygen saturation and diameter, and are used to estimate the LoA for the two versions. As there are multiple observations per image the LoAs are calculated for multiple replicates with 95% CIs calculated using the MOVER method [25]. Finally, the time in seconds taken to analyze an image is measured for both versions. The mean and standard deviation are calculated.

Statistical significance of the difference is evaluated using a two-tailed, paired t-test. Statistical analysis and generation of plots and graphs was performed in R version 3.6.1 [26] using irrICC [27] version 1.0, ggplot2 [28] version 3.3.2 and boot [29, 30] version 1.3-25.

## Results

Figure 2 shows saturation and diameter measurements for the first image of each patient. Oxygen saturation in retinal arterioles was different between analysis software; 93.7%±3.3% for the automatic version 3.0 and 90.7%±3.8% for the semi-automatic version 2.5 (*p <* 0.0001, mean±SD). Venular oxygen saturation was lower measured with version 2.5, 58.9% ±5.1% compared to version 3.0; 62.3% ±5.8%, (*p <* 0.0001, mean±SD). Standard deviation between measured subjects was lower in version 3.0, compared to version 2.5 for arterioles (*F* = 1.47, *p* = 0.04) but the difference was statistically insignificant for venules (*F* = 0.74, *p* = 0.12). When diameter measurements where compared between analysis software, arteriolar diameter was decreased with the new version compared to the old version (9.7 ± 1.1 pixels vs. 12.3±1.1 pixels; *p <* 0.0001) as well as the venular diameter (12.9±1.4 pixels vs. 15.5±1.0 pixels; *p <* 0.0001). The standard deviation of arteriolar diameter measurements was not significantly altered (*F* = 1.22, *p* = 0.14) but standard deviation of venular measurements was significantly less for version 3.0 compared to version 2.5 (*F* = 1.83, *p <* 0.0001).

**Fig 2.**
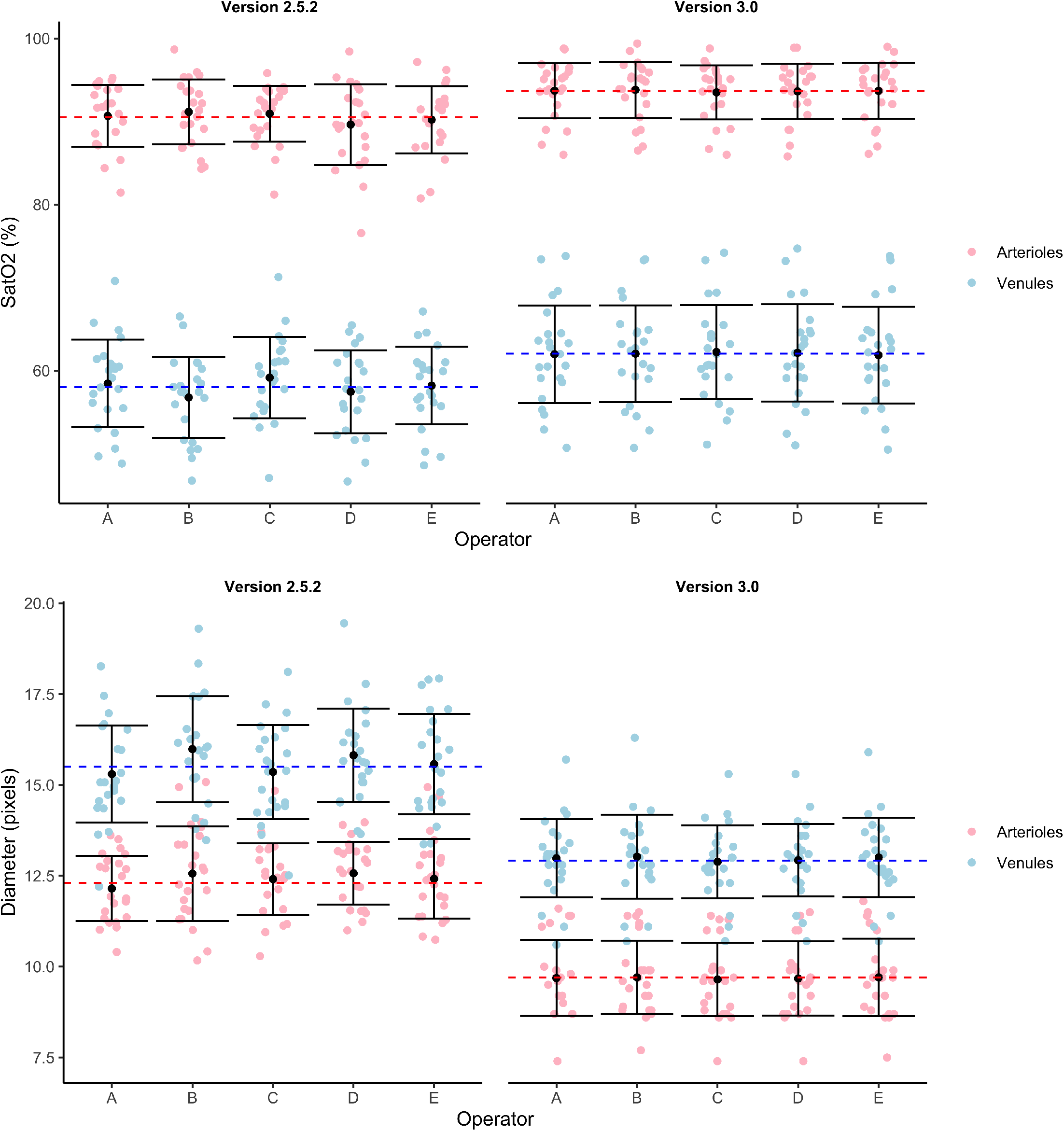
Measured vessel saturation (top) and diameter (bottom) values from the first image for the two versions and all operators. Light red and blue points show individual measurement values, dashed red and blue lines show the global mean values for arterioles and venules, respectively. Black dots show the mean, and error bars show the standard deviation around the mean for each operator and vessel type.

Figure 3 shows Bland-Altman plots for saturation and diameter measurements.

**Fig 3.**
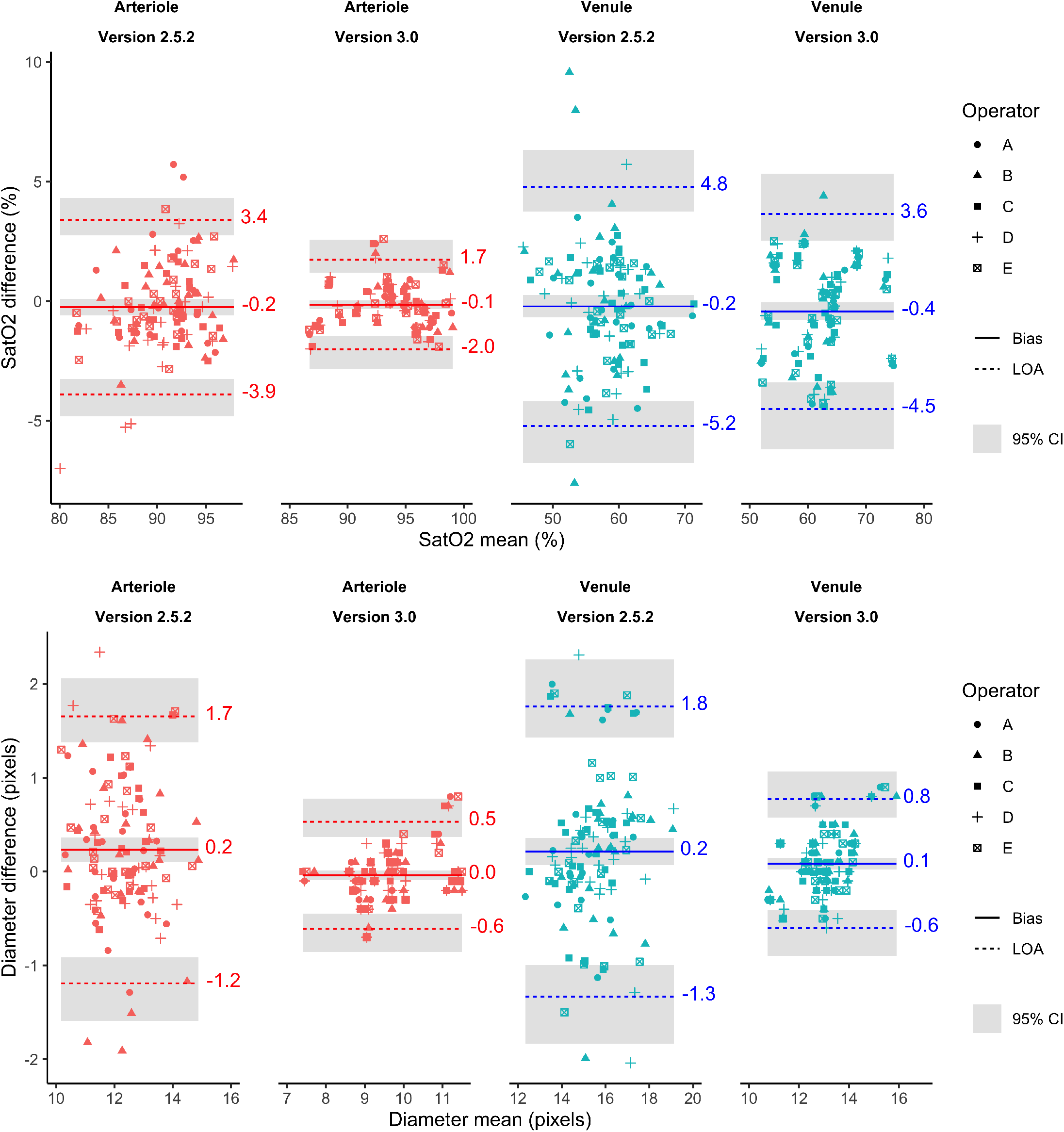
Analysis of repeatability of retinal vessel oximetry (top) and diameter (bottom). The same area in two images acquired within a short time interval is measured using the two software versions for arterioles and venules. The vertical axis shows the difference between two saturation and diameter measurements in the same area (first image minus second image), and the horizontal axis shows their mean. The solid line in the middle shows the mean difference, and the other two broken lines show Limits of Agreement (LoA) or the two standard deviations of the difference. Results are colored red for arterioles and blue for venules. Grey shaded areas show the 95% confidence interval around the bias and LoA.

Table 1 shows intra and inter-rater reliability when measuring vessel oxygen saturation.

**Table 1.** Intra and inter-rater reliability (ICC) when measuring vessel oxygen saturation using the two versions. Measurements from the same area of 23 right eyes in 23 subjects.

|  | Version | ICC<sub>intra</sub> [95% CI] | ICC<sub>inter</sub> [95% CI] |
| --- | --- | --- | --- |
| Arterioles | 2.5 | 0.80 [0.70, 0.90] | 0.81 [0.71, 0.90] |
| Arterioles | 3.0 | 0.97 [0.95, 0.99] | 0.97 [0.95, 0.99] |
| Venules | 2.5 | 0.80 [0.68, 0.90] | 0.84 [0.75, 0.92] |
| Venules | 3.0 | 0.96 [0.94, 0.98] | 0.96 [0.94, 0.98] |

For arterioles measured with version 2.5 the intra and inter-rater reliability is moderate to good [22]. For venules measured with version 2.5 the intra-rater reliability is moderate to good and the inter-rater reliability is good to excellent. For arterioles and venules measured with version 3.0 the intra and inter-rater reliability is excellent.

Table 2 shows the intra and inter-rater reliability when measuring vessel diameter. For arterioles and venules measured with version 2.5 the intra and inter-rater reliability is moderate to good. For arterioles and venules measured with version 3.0 the intra and inter-rater reliability is excellent.

**Table 2.** Intra and inter-rater reliability (ICC) when measuring vessel diameter using the two versions. Measurements from the same area of 23 right eyes in 23 subjects.

|  | Version | ICC<sub>intra</sub> [95% CI] | ICC<sub>inter</sub> [95% CI] |
| --- | --- | --- | --- |
| Arterioles | 2.5 | 0.71 [0.58, 0.84] | 0.73 [0.60, 0.85] |
| Arterioles | 3.0 | 0.97 [0.96, 0.99] | 0.97 [0.96, 0.99] |
| Venules | 2.5 | 0.71 [0.56, 0.84] | 0.76 [0.65, 0.87] |
| Venules | 3.0 | 0.95 [0.91, 0.98] | 0.95 [0.92, 0.98] |

The analysis time per image was 617 ± 187 seconds for version 2.5 and 29 ± 7 seconds for version 3. Using version 3.0 the analysis of images was significantly faster than for version 2.5 (p = 0.032).

## Discussion and conclusions

This study compares two versions of software capable of measuring retinal vessel oxygen saturation and vessel diameters. Vessels from the same area were measured in two images taken of the same subject with a short time interval. One version (v3.0) automatically selects the vessels to be measured and classifies them as arterioles or venules, while the other version (v2.5) requires operator input for these tasks.

Version 3.0 measures higher oxygen saturation values for both arterioles and venules than version 2.5 but the variability between measured subjects is similar or slightly lower for version 3.0.

The limits of agreement analysis for oxygen saturation indicate that repeatability is substantially improved for arterioles but less so for venules. The intra and inter-rater ICCs indicate that repeatability, and agreement between operators is improved for version 3.0 compared with version 2.5.

When measuring vessel diameter version, 3.0 performs substantially better for both arterioles and venules compared with version 2.5. The lower measured diameter in version 3.0 versus version 2.5 can be attributed to superior vessel detection where the boundaries of vessels are more precisely located. Measurements with version 3.0 therefore include some of the smaller vessels that were not detected by version 2.5. As this might lead one to suspect that the lower LoA was simply an artifact of these lower measurement values, the diameter measurements were also compared after the values from version 3.0 had been scaled to have the same mean value as the diameter measurements from version 2.5. Even after this adjustment, the widest LoA for version 3.0, taking into account the outer limits of the 95%CI, was narrower than the narrowest LoA for version 2.5, where the inner limits of the 95% CI were used. We, therefore, conclude that there is substantial evidence that the diameter measurements for version 3.0 are more repeatable than for version 2.5.

This study has several limitations which should be addressed in follow up studies. Here, only images from young, healthy volunteers were used. While anecdotal evidence shows that the automated software works well with images from patients suffering from conditions such as CRVO and diabetic retinopathy the performance should be validated beyond the healthy eye.

In conclusion, while both versions of the software evaluate the oxygen saturation in a similar manner, the measured values are slightly different, which means that results from the two software versions are not directly comparable. The automatic software (Version 3.0) gives more consistent results, both for repeated measurements and between operators, than the manual software (version 2.5). Finally, the automated measurements are substantially faster than the manual measurements.

## Data Availability

All data is available in the manuscript.

## 1 Acknowledgments

